# An exploration of methods to enable equitable access to non-invasive prenatal screening

**DOI:** 10.1101/2022.06.17.22276572

**Authors:** Mathias Ehrich, Katelynn G. Sagaser, Christopher K. Ellison, Allison Chia-Yi Wu, Don C. Hutchison, Richard P. Porreco, Deborah Bellesheim, Avinash S. Patil, Lee P. Shulman, Dirk van den Boom

## Abstract

**Objective:** To explore capillary blood collection as a potential method to enable more equitable access to non-invasive prenatal screening (NIPS) for aneuploidy.

**Methods:** All participants contributed venous and capillary blood samples in this comparative study. Samples were analyzed using standard NIPS methods. Venous samples were used as the basis. Multiple parameters were compared including specimen collection, cfDNA characteristics, and NIPS outcome.

**Results:** Venous and capillary sample pairs were successfully collected for comparative analysis from 202 participants. Cell free DNA (cfDNA) size profiles from venous vs capillary blood were not different (*p* >0.05). Evaluation of fetoplacental cfDNA contribution in plasma revealed no statistically significant difference in venous vs capillary samples. Elevated Z-scores for trisomy 21 (n=13), trisomy 18 (n=2), or trisomy 13 (n=1) were concordant between venous and capillary samples in all 16 pairs.

**Conclusion:** We have demonstrated that sufficient capillary blood volumes can be obtained for NIPS. Furthermore, capillary and venous cfDNA samples have comparable characteristics. As NIPS results from capillary blood collections are equivalent to NIPS results obtained from venous blood, capillary blood collections are a potential candidate for a distributable NIPS system which promotes equitable access to NIPS for all pregnant patients.

## INTRODUCTION

Prenatal screening has been fundamentally changed within the last decade by the introduction of circulating cell free DNA (cfDNA) analysis. While initially met with hesitancy, non-invasive prenatal screening (NIPS) for fetal aneuploidy using cfDNA analysis is now widely accepted in the obstetrical community.^1,2^ Despite great technical success and recent endorsements for use in all pregnancies,^1-3^ universal adoption of NIPS remains hindered by various barriers that undermine equitable NIPS access.

Several innovative yet unsuccessful attempts have been made to offer lower cost NIPS, each aimed at changing the detection technology from next generation sequencing to methods like RT-PCR, high molecular counting, or single molecule detection.^4-7^ Our experience at a large commercial NIPS laboratory demonstrated that elements such as sample logistics and overhead costs can account for as much as 90% of total cost.

To mitigate NIPS access barriers, a solution must be both distributable and feasible for self-collection. Circulating cfDNA can be detected in different types of self-sampled biological specimens ; yet, the practical application of cfDNA analysis varies by sample type. Extracted cfDNA from urine is too short for NIPS,^8^ and reliable cfDNA collection from sputum and sweat has not been possible in our initial tests (data not shown).

Self-collected capillary blood has been successfully utilized for fetal sex prediction through Y chromosome analysis.^9-11^ In this study, we explore capillary blood collection as a possible method to enable more equitable access to NIPS. We compare NIPS results from capillary blood to results from venous blood.

## METHODS

### Blood collection

Sample collection was executed in two separate study arms through a study protocol approved by the Aspire Independent Review Board (IRB #Juno-2017-0001). In both study arms, venous blood was obtained through traditional venipuncture. A trained medical professional (e.g., nurse, phlebotomist, or physician) performed the capillary blood collection in the assisted collection study arm. In the self-collection study arm, the participant was provided with self-collection instructions and independently collected a capillary blood sample.

Blood was processed to plasma by a standard centrifugation protocol and the resulting plasma was stored at -80C until time of analysis.

### NIPS analysis

A protocol for NIPS was established based on previously published, well-described methods.^12^ Sequencing was performed on the Illumina NextSeq550 and NextSeq2000 platforms. The resulting sequencing data were analyzed using bowtie2 for sequence alignment, and trisomy classification was performed according to previously described methods.^13^

Data analysis was performed in the R statistical package^14^ using a data analysis method validated in a set of >1,200 maternal samples, including 326 samples with NIPS results positive for fetal trisomies. These samples were split into a training set (n=671, including 84 trisomy 21 samples, 74 trisomy 18 samples, and 21 trisomy 13 samples) and a test set (n=550, including 75 trisomy 21 samples, 56 trisomy 18 samples, and 16 trisomy 13 samples). Calling routines and thresholds were established using the training set data. The test set data was signed out by a laboratory director blinded to the outcome. Sensitivity and specificity were established based on the test set data and each exceeded 99%.

## RESULTS

### Sample collection

A total of 207 venous and capillary sample pairs were successfully collected. Of 414 individual samples, two venous and three capillary samples failed quality control and therefore were excluded from analysis. As all samples were acquired as pairs, a total of 202 sample pairs were available for comparative analysis, offering a first pass rate of >98.5%.

Between assisted collection samples and self-collected samples, no statistically significant differences were observed in the clinically relevant data for weight, height, maternal age, and gestational age (Table 1). Gestational age showed a slightly different distribution across the two groups (Figure 1). Samples collected in the assisted study arm demonstrated a bimodal distribution clustered with the timing of those in-office visits which typically follow the identification of abnormal findings on first trimester screening and second trimester anatomic survey. In contrast, samples in the self-collected arm were more uniformly distributed across gestational ages.

**Table 1:** Participant clinical characteristics by sample collection method

|  | Assisted Collection,<br>n = 96<br>Mean value (SD) | Self-Collection,<br>n = 106<br>Mean value (SD) | *p*-value (statistical analysis) |
| --- | --- | --- | --- |
| Age (years) | 29.7 (8.1) | 32 (5.4) | 0.4 (t-test) |
| Weight (kg) | 76.8 (15.4) | 80.2 (21.4) | 0.2 (t-test) |
| Height (m) | 1.66 (0.08) | 1.64 (0.07) | 0.02 (t-test) |
| BMI (kg/m<sup>2</sup>) | 27.9 (5.3) | 29.7 (7.1) | 0.05 (t-test) |
| Gestational<br>Age (weeks) | 23.4 (7.5) | 21.7 (9.1) | 0.1 (Wilcoxon rank sum test) |

**Figure 1:**
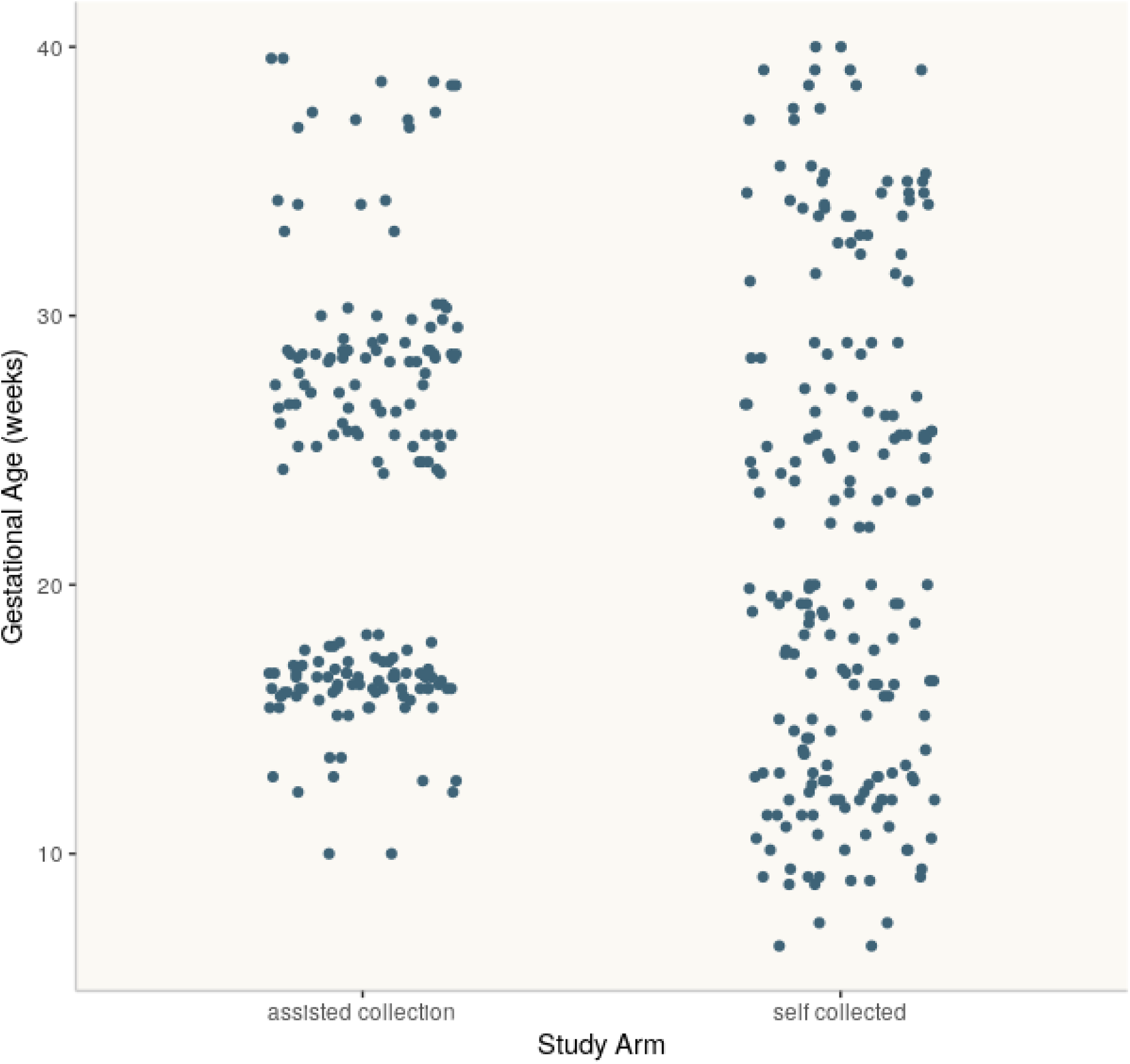
Distribution of participant gestational ages at time of capillary sample self-collection vs assisted collection

### Size profile

Fragment length profiles for venous cfDNA are well established with a maximum around 168bp and a periodicity in 10bp intervals.^15,16^ Scarcity of shorter DNA fragments is typically associated with lower contributions of fetoplacental DNA. Size profiles for cfDNA from capillary blood and venous blood were largely overlapping, and no statistically significant difference was identified over the entire profile (Wilcoxon paired rank test, *p*>0.05) (Figure 2).

**Figure 2:**
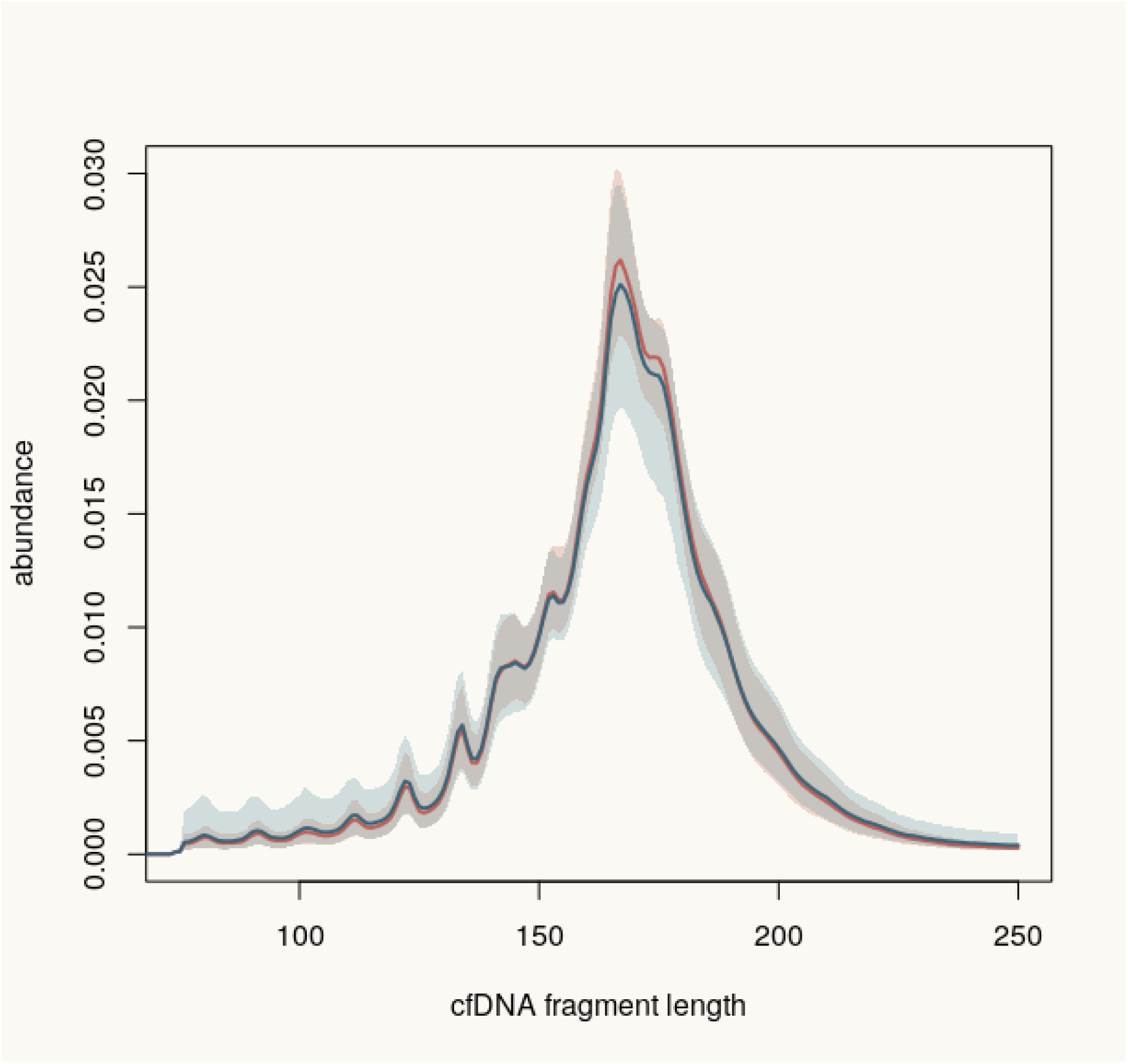
Cell free DNA fragment length profiles in capillary blood and venous blood samples

### Venous to capillary comparison

#### Y chromosome

Evaluation of Y chromosome contribution, an indicator of fetoplacental cfDNA contribution in plasma, revealed no statistically significant difference in samples from capillary blood and those from venous blood. Samples with Y chromosome representation above a threshold of 6e-5 can be assumed to contain cfDNA from a pregnancy with at least one Y chromosome, as is typically observed with a male fetus. In contrast, samples with Y chromosome representation below 5e-5 can be assumed to not have Y chromosome DNA, as is typically observed with a female fetus.

Results were concordant for the Y chromosome between venous and capillary blood samples in 198 sample pairs (Figure 3). Four samples demonstrated discordance, with two samples showing Y chromosome DNA only in capillary samples and two showing Y chromosome DNA only in venous samples. Reanalysis of these four samples revealed concordance between the capillary and venous sample pairs for two of the pairs, suggesting that the discordance is not a feature of the sample itself but rather a consequence of the stochastic effects during NIPS.

**Figure 3:**
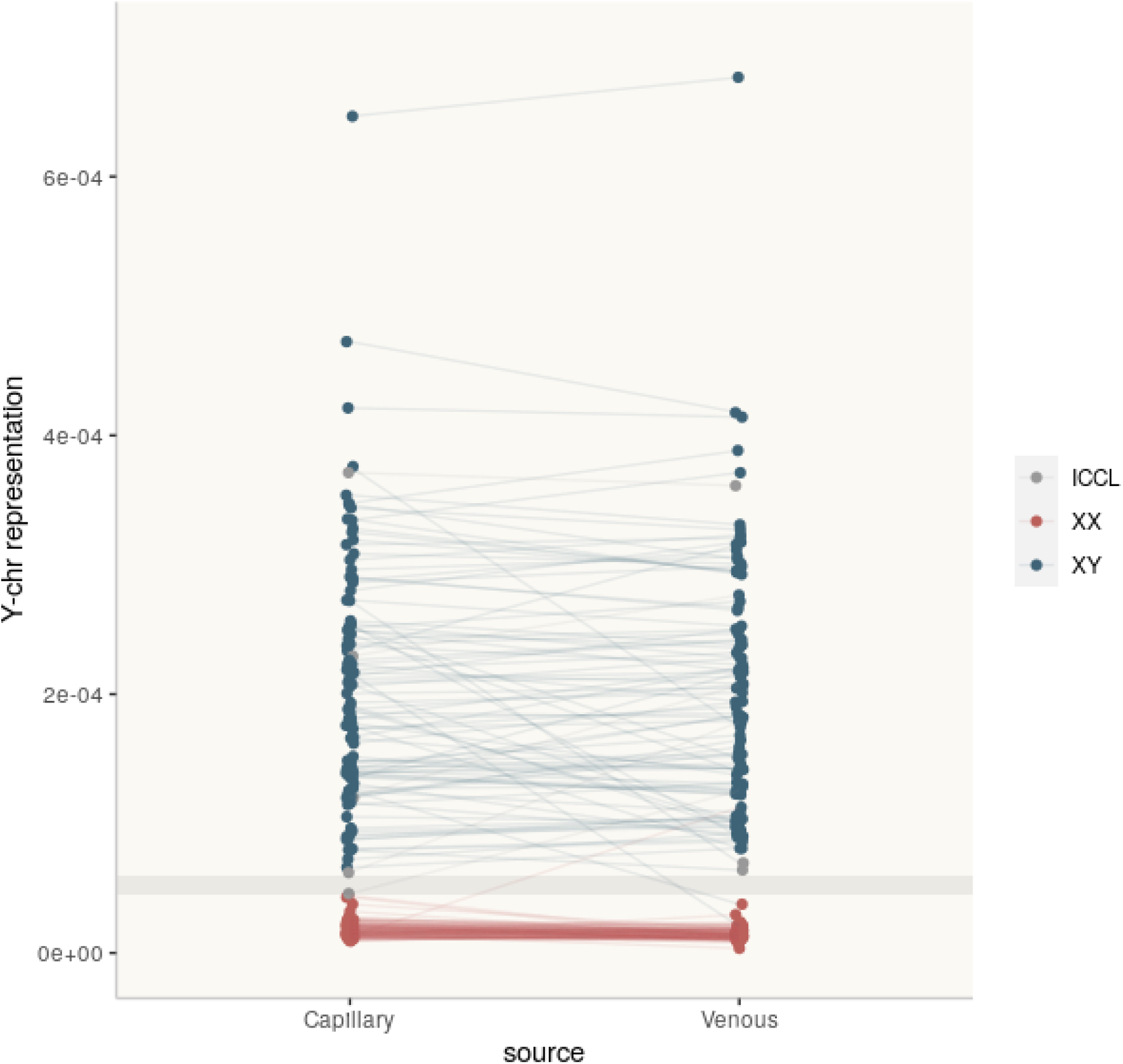
Y chromosome representation across capillary and venous samples. Inconclusive (ICCL); Consistent with female (XX); Consistent with male (XY)

#### Trisomies

Elevated Z-scores for one of the three common autosomal trisomies were observed in 16/202 (7.9%) sample pairs (trisomy 13, n = 1; trisomy 18, n = 2; trisomy 21, n = 13) (Figure 4). Venous and capillary result concordance was observed in all 16 pairs with elevated Z-scores for common trisomies. An inconclusive result between capillary or venous pairs was generated for 2/202 sample pairs (1.0%). Specifically, one sample was euploid in the venous sample and inconclusive in the capillary sample, while the other was euploid in the capillary sample and inconclusive in the venous sample. Reanalysis of inconclusive results demonstrated concordant euploid results for capillary and venous samples, suggesting the differences are likely attributable to stochastic variation.

**Figure 4:**
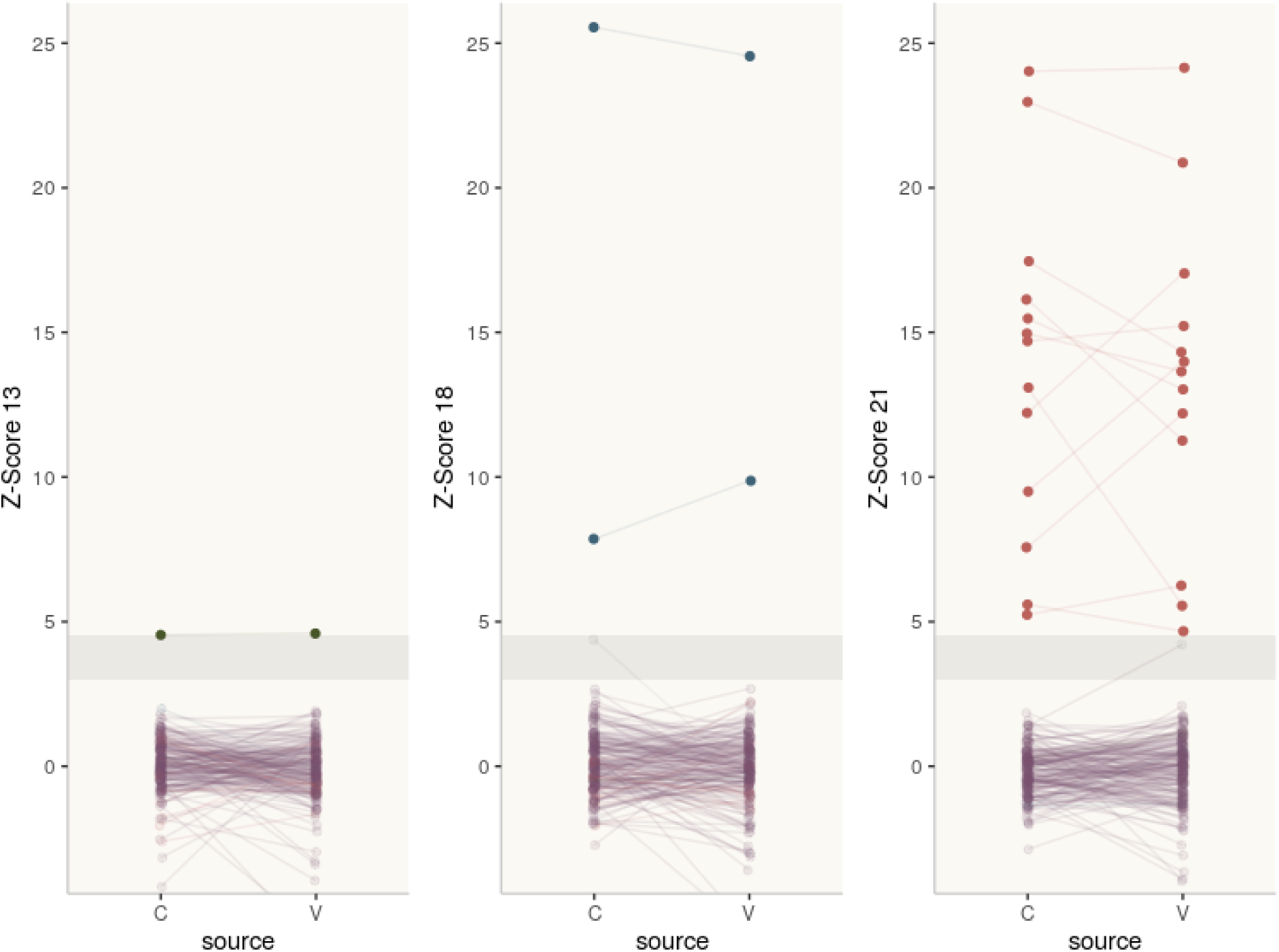
Z-scores for common autosomal trisomies across capillary and venous samples. Capillary (C); Venous (V)

#### Unexpected findings

When applying a comparative study design, diagnostic outcomes are not necessary for all samples; however, our study included a subset of samples with screening outcomes available at the study onset. One sample with available screening results was reported to be high risk for trisomy 13 by traditional NIPS. Our results of both capillary and venous material indicated a euploid result which was confirmed after delivery. Furthermore, three samples were externally flagged as “high risk for trisomy 13 and trisomy 18.” In all three of these samples, our results indicated euploidy within capillary and venous material and provider follow-up confirmed our results.

## DISCUSSION

Enabling widespread, equitable access to NIPS requires an approach that is affordable and attainable without additional office visits. Cell free DNA remains the most accurate biomarker for NIPS but has only been performed from venous collection. Our reimagined vision of the NIPS paradigm investigated capillary blood as an input material for NIPS. After establishing a laboratory workflow compatible with low input volumes, we focused on the collection and evaluation of capillary blood. We successfully demonstrated three elements crucial to the potential implementation of a more equitable NIPS model using capillary samples. First, sufficient amounts of capillary blood can be obtained with or without assistance from medical personnel. Second, cfDNA from capillary blood has comparable characteristics as cfDNA from venous blood. Third, NIPS results from capillary blood collections are equivalent to NIPS results obtained from venous blood. Therefore, capillary blood collections are a prime candidate to enable and build a distributable NIPS system.

To enable a comprehensive implementation of NIPS as a screening tool for all pregnancies, the issues of NIPS cost and access must each be improved. Half of all counties in the United States lack an OB/GYN, leaving 10 million birthing people with little to no obstetrical care.^17,18^ Access to U.S. prenatal care is unevenly distributed and primarily driven by income, location, and insurance status. Only 21 states’ Medicaid programs offer NIPS coverage for all pregnant people.^19^ A fragmented health insurance landscape and questionable laboratory billing practices have created enormous difficulties for patients seeking an accurate estimate of their NIPS out-of-pocket responsibilities.^20^

The dilemma of logistical access to NIPS in the clinic or at the lab is no small matter. Today, all NIPS assays require a venous blood draw and, consequently, a phlebotomy appointment. For patients with limited access to care, minimally flexible schedules due to child-or elder-care considerations, or limited time away from work, any office visit can be a physical and/or financial challenge. Research exploring ambulatory medical care opportunity costs identified a two hour average in-person appointment time.^21^ The average lost wages associated with seeking in-person care was $47, higher than the average co-pay for care.^21^ Innovations in the delivery of accessible prenatal care include the 4-1-4 prenatal plan^22^ in which average-risk patients may tailor their care beyond the standard of 4 in-person visits, 1 anatomy ultrasound visit, and 4 virtual visits. Even with the 4-1-4 prenatal plan’s conveniences, implementation of first trimester NIPS into routine care would require an additional in-person visit for phlebotomy given in-person visit spacing.

Capillary blood collection methods for NIPS allow the flexibility of sample collection in the office or at home. While sufficient amounts of capillary blood were obtained for every participant in our study, venous blood collection was not successful in four participants. Our protocol limited a maximum of two venous draw attempts, and all collections were performed by experienced, licensed phlebotomists. Difficult venous access is a frequently encountered phenomenon most commonly addressed by scheduling a subsequent phlebotomy appointment that may also involve further difficulties in venous access. The option of a fingerstick performed by a friend or office staff would be of great benefit in a larger population inclusive of individuals with an aversion to self-collection – a behavior not observed in our study.

A dispersed NIPS system centered on equitable and comprehensive access to the genetic information afforded by NIPS requires careful consideration of an accompanying platform for counseling information and support. Such a platform should ensure successful sample collection for all patients, and particularly those with low health literacy levels, limited familiarity with biological sample collection, and differing abilities. This support might include materials such as video content, live support, or in-person assistance. In addition, available resources should include engaging educational content to support pre- and post-test genetic counseling for NIPS, as well as genetic counseling services in real time via telehealth, as counseling availability is critical for effective prenatal screening programs.

Without traditional phlebotomy requirements, a dispersible self-collection system for NIPS offers the potential to better serve patients. Rather than replacing the indispensable role of the clinician, at-home access to NIPS and accompanying information can complement traditional office visits and telehealth offerings alike. In so doing, clinicians providing OB/GYN care may benefit from more streamlined, patient-centered workflows without laboratory logistical concerns. Future work should continue to evaluate the accuracy of capillary blood collection methods in a larger sample cohort as well as explore the socioeconomic opportunities a dispersible NIPS system may provide.

## Data Availability

All data produced in the present study are available upon reasonable request to the authors

## Notes

### Competing Interest Statement

Authors ME, KGS, CKE, ACYW, DCH, and DvdB are employees and shareholders of Juno Diagnostics. Author DB receives an honorarium for service on Juno Diagnostics clinical advisory board.

### Funding Statement

This study did not receive any funding

### Author Declarations

Aspire Independent Review Board gave approval for this work (#Juno-2017-0001).

